# Cisplatin promotes pyroptosis of gastric cancer cells by activating GSDME

**DOI:** 10.1101/2023.09.08.23295232

**Authors:** Xianglai Jiang, Yongfeng Wang, Chenyu Wang, Haizhong Ma, Miao Yu, Hui Cai

## Abstract

According to studies, numerous chemotherapeutic drugs can facilitate programmed cell death via pyroptosis. Clarifying the mechanism by which cisplatin kills gastric cancer cells is crucial for enhancing gastric cancer’s sensitivity to chemotherapy and elucidating the mechanism of drug resistance in gastric cancer. The differentially expressed genes following cisplatin treatment were identified using second-generation sequencing technology. Bioinformatics was used to investigate the functional enrichment of differentially expressed genes and core genes in tumor cells killed by cisplatin. Cox regression analyses were used to examine the pyroptosis core genes that worked as independent prognostic factors for patients with gastric cancer. The expression of core genes in gastric cancer cells was silenced by siRNA, and the changes in the proliferation of gastric cancer cells were observed. The expression of related genes and the survival of gastric cancer cells after the addition of cisplatin were observed. The second-generation sequencing, RT-PCR and Western blotting showed that the pyroptosis core gene was significantly highly expressed after cisplatin treatment. The results of differential gene enrichment of cisplatin-treated gastric cancer cells showed that differential genes were mainly concentrated in biological processes and signaling pathways related to pyroptosis. GSDME protein is highly expressed after cisplatin treatment, and it is also a poor prognostic factor for gastric cancer patients and an independent prognostic factor. After the same dose of cisplatin treatment, the survival rate of siGSDME gastric cancer cells was significantly higher than that of GSDME regular expression gastric cancer cells. After acting on gastric cancer cells, cisplatin triggers pyroptosis by stimulating the activation of genes such as GSDME, resulting in the death of gastric cancer cells. GSDME is an independent prognostic factor for gastric cancer patients and is significantly linked with a shorter OS. In gastric cancer cells, silencing GSDME can substantially reduce cisplatin’s cytotoxicity.

## Introduction

Gastric cancer is the fourth leading cause of cancer death (7.7 %), with more than 700,000 deaths worldwide in 2020^1–4^. The origin of gastric cancer can be any part of the stomach and can spread to the whole stomach and even other organs (such as colorectal, liver and ovary)^1^. Gastric cancer is a disease with high molecular and phenotypic heterogeneity: in a multi-stage cascade process, gastritis caused by chronic Helicobacter pylori eventually develops into gastric cancer after precancerous stages of atrophic gastritis, intestinal metaplasia and dysplasia ^5^. The mechanism of gastric cancer is complex and diverse, the biological processes involved include somatic mutation, mutation of proto-oncogenes and tumor suppressor genes, pyroptosis, apoptosis, and oxidative damage of DNA^6^. The initial symptoms of gastric cancer are not obvious, and clinical signs often manifest in the advanced stage of cancer^7^. When gastric cancer progresses to an unresectable stage, chemoradiotherapy is the main treatment^8^. In treating gastric cancer patients, the emergence of drug resistance and metastasis often means the failure of treatment. Therefore, it is significant to study the mechanism of gastric cancer chemotherapy and put forward new ideas to block the generation of drug resistance and improve sensitivity. ^9, 10^.

Cells are the basic unit of life activities, and cell death, as a natural life phenomenon, is significant for maintaining the relative stability of the body’s internal environment of the body^11^. Pyroptosis and apoptosis are both programmed cell death, and apoptosis will shrink into apoptotic bodies and be swallowed by phagocytes, so it does not cause inflammatory response^12^. The concept of pyroptosis was first proposed by Cookson and Brennan in 2001 to describe a cysteine protease 1 (CASP1) -dependent cell death in inflammatory cells^13^. Cells undergoing pyroptosis continue to expand until the cell membrane ruptures, leading to the release of intracellular substances and the activation of a robust inflammatory response^14^. Pyroptosis is an indispensable natural immune response mediated by Gasdermin protein in the human body^14^. After receiving inflammatory stimulation, caspase family proteins cut Gasdermin protein and cause the cut Gasdermin protein to aggregate on the surface of the cell membrane to form a cavity, to achieve the role of cell content material outflow and cell death^15^. As an essential protective mechanism of human body, pyroptosis can timely remove infected or mutated abnormal cells and the presence of pyroptosis has an important anti-tumor effect^16^. Cisplatin is a platinum-containing anticancer drug. It is an orange or yellow crystalline powder, slightly soluble in water, easily soluble in dimethylformamide, and can be gradually converted and hydrolyzed in an aqueous solution^17, 18^. Clinical use of cisplatin in ovarian cancer, prostate cancer, testicular cancer, lung cancer, nasopharyngeal carcinoma, esophageal cancer, malignant lymphoma, head and neck squamous cell carcinoma, thyroid cancer and osteosarcoma and other solid tumors can show efficacy^19, 20^. Recent studies have shown that cisplatin can activate pyroptosis to mediate gastric cancer cells after acting on cancer cells, which has a new direction for the mechanism of cisplatin as a chemotherapeutic drug to kill gastric cancer cells^21–23^.

In this study, basic experimental techniques and bioinformatics analysis methods were used to identify the significantly differentially expressed apoptosis core genes and pyroptosis core genes after cisplatin treatment of gastric cancer cells. The expression of GSDME, a highly expressed pyroptosis execution factor after cisplatin treatment, in gastric cancer and its effect on the biological function of gastric cancer cells were explored. Finally, it was confirmed that silencing the expression of GSDME in gastric cancer cells reduced the sensitivity of gastric cancer cells to cisplatin, which proved that cisplatin mediated pyroptosis of gastric cancer cells by activating GSDME. It was found that GSDME was an independent prognostic factor for gastric cancer patients, and could potentially be a prognostic marker, chemotherapeutic drug sensitivity and immunotherapy biomarker.

## Material and method

### 2.1 Cell culture

Human gastric epithelial cell (GES-1), human gastric adenocarcinoma cell (AGS) and human poorly differentiated gastric cancer cell (MKN45) were purchased from the Cell Resource Center of Shanghai Academy of Sciences. The cells were cultured in 4ml RPMI-1640 medium containing 10 % fetal bovine serum (Biological Industries, Israel) and placed in a 37 °C constant temperature cell incubator containing 5 % CO2. All cells were subcultures when the growth length reached 80 %, and all cell lines maintained the typical morphology of the cells during the study.

### 2.2 Determination of semi-inhibitory concentration

AGS cells and MKN45 cells without abnormal morphology and growth status were obtained. After the cells reached the preset concentration and were in the logarithmic phase of growth observed by the microscope, cisplatin (APExBIO, USA) of 0μM, 5μM, 12.5μM, 25μM, 50μM and 100μM were added respectively, and the cells were transferred to 37 °C and 5 % CO2 cell incubator for 12 h. After 12 hours of culture, 100ul of the mixture was gently removed and 10ul of CCK8 reagent (APExBIO, USA) and 100ul of serum-free medium (Biological Industries, Israel) were added. The cells were cultured in a 37 °C thermostatic shaker for 1 hour, and then the OD value of each well was measured at a wavelength of 450 nm on a microplate reader (Thermo Scientific, USA). The measured OD value was converted into a percentage value to obtain the drug concentration and the corresponding inhibition rate and imported into EXCEL software and GraphPad Prism 8.0 software. The IC50 of cisplatin on gastric cancer cells AGS and MKN45 for 12 hours was calculated and the corresponding figures were drawn.

### 2.3 The second-generation sequencing detection

MKN45 cells were treated with IC50 cisplatin, and the total RNA of MKN45 cells was extracted by Invitrogen TRIzol as the initial input material. The initial input material is enzymatically shredded to conform to the fragments required by the Illumina machine. The single-stranded DNA fragment in the sample was entered into the flow cell and then combined with the P5 ’ of the flow cell to complete the bridge PCR amplification. A special dNTP with four different fluorescent groups was added to emit different fluorescence signals at the same time, and then the extension-scanning process was repeated until the entire DNA sequence was spliced. The differentially expressed mRNAs before and after gastric cancer cell MKN45 were sequenced using the principle of reversible termination of chemical reactions through four special deoxyribonucleotides that can emit fluorescence signals.

### 2.4 Real-time fluorescent quantitative PCR

The total RNA of 10 pairs of gastric cancer clinical samples was extracted strictly according to the manufacturer’s requirements. The extracted RNA was reverse transcribed into cDNA using a reverse transcription kit (vazyme, China), and the reaction system was configured according to 10ul SYBR green reagent, 7.2ul DEPC water, 2ul cDNA, 0.4ul pre-primer and 0.4ul post-primer. According to the pre-deformation: 95 °C / 5 minutes (one cycle) ; cyclic reaction : 95 °C / 10s-60 °C / 30s (40 cycles) ; dissolution : 95 °C / 15s-60 °C / 60s-95 °C / 15s (one cycle) reaction procedure for PCR quantification. The quantitative fluorescence analysis of GSDME with GAPDH as the internal reference gene was completed under the lighter machine (Roche, Switzerland). The expression of GSDME in different tissues was calculated according to the 2*^-ΔΔCT^* method, the statistical analysis was completed in GraphPad Prism 8.0 software, and the corresponding patterns were drawn.

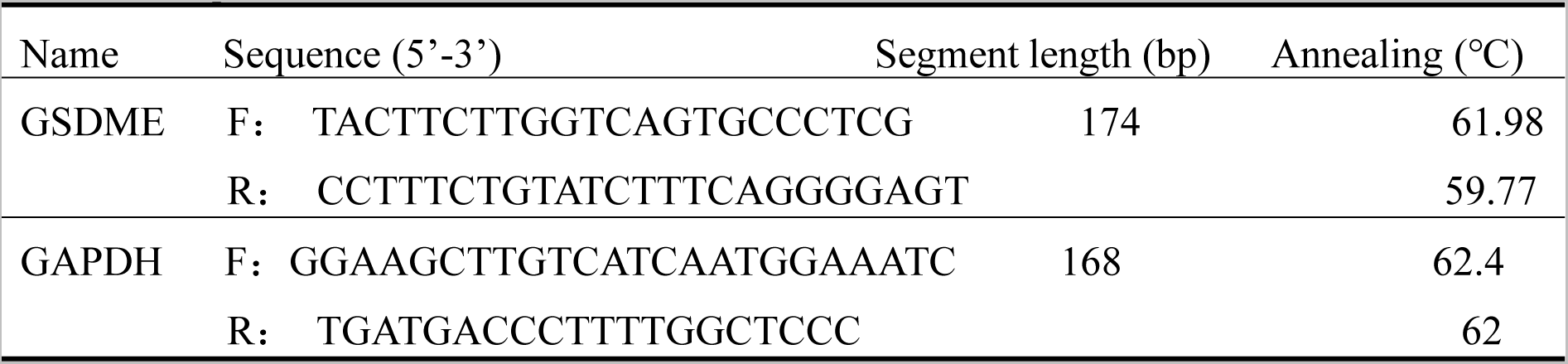
Primers Sequences.

### 2.5 Western blot detection

The treated GES-1, AGS and MKN45, as well as MKN45 and AGS co-cultured with cisplatin for 24 h were taken out in an incubator at 37 °C, and the proteins were extracted and quantified. The protein separated by electrophoresis was transferred to the PVDF membrane, quickly blocked for 30 minutes, then placed in a specific primary antibody (abcam, UK) and incubated in a 4 % environment for 12 hours. Then the anti-rabbit antibody (proteintech, China) was incubated at room temperature for 2 hours, and the gel imaging system (eBlot, China) was used for imaging after using the chromogenic agent.

### 2.6 mRNA data collection

The RNA-Seq expression profile data of TCGA-STAD were obtained by logging into the TCGA database website (https://www.cancer.gov/) in December 2022. At the same time, the expression profiles of cancer tissues and adjacent tissues of gastric cancer patients and the clinical information of gastric cancer patients were collected from the TCGA database. The expression of core genes in gastric cancer was analyzed using the GEPIA 2 website (http://gepia2.cancer-pku.cn/) in December 2022^24^. The immunohistochemical images of core gene protein expression in ’ stomach ’ and ’ gastric cancer ’ were downloaded from the human protein atlas (https://www.proteinatlas.org) database in January 2023.

### 2.7 Single-factor regression analysis

To prepare the correlation expression of core genes in stomach adenocarcinoma (STAD) samples in each TCGA database and the corresponding survival status and survival time matrix, as well as the matrix that converts clinical information (age, gender, grade, stage staging, T staging, M staging and N staging) into continuous variables. The prepared matrix information was input, and the ’ bioForest ’ function and ’ indep ’ function in the ’ survival ’ software package in R software were applied to complete the single factor Cox regression analysis of core genes for gastric cancer patients to explore whether core genes can be used as independent prognostic factors for gastric cancer patients. The expression data of ten pyroptosis-related genes CASP1, CASP8, CHMP6, GSDMD, GSDME, IL6, NCRC5, NLRP1, NLRP6 and TP53 highly expressed after cisplatin stimulation were selected as continuous variables in the Cox regression model for analysis.

### 2.8 Ethics

This study was approved by the Medical Ethics Association of Gansu Provincial People ’s Hospital, ethical number: 2022316. All patients signed the informed consent form and agreed to the study, and all processes were in line with the Declaration of Herkissin. After the patient ’s family members and patients have been informed and obtained their permission, the abandoned cancer tissues and adjacent tissues of patients who underwent gastric cancer resection in Gansu Provincial People ’s Hospital from August 28, 2022 - September 30, 2022 were collected, which conforms to the principle of controlling risks and protecting the safety and health rights of subjects. RNA was extracted from discarded tumor tissues and adjacent tissues and stored in a refrigerator at -80 °C. The information of all subjects was strictly confidential, and the relevant information was not disclosed to any third party without the authorization of the subject, in line with the principle of privacy protection.

### 2.9 Gene enrichment analysis

The analysis included different genes with p values less than 0.05 before and after cisplatin treatment. Gene ontology (GO) and Kyoto Encyclopedia of Genes and Genomes (KEGG) enrichment analysis were performed on the differential gene set using the ’org.Hs.eg.db ’R software package^25^. According to the expression level of gastric cancer patients in the TCGA database, they were divided into two groups, and the differentially expressed genes in the core gene high expression group and the core gene low expression group were identified. The differential genes of the two groups of core genes were analyzed by GO and KEGG enrichment analysis.

### 2.10 Kaplan-Meier survival analysis

According to the median value of core gene expression, gastric cancer patients in the TCGA database were divided into high core and low core gene expression groups. The comparison of OS between the two groups with high core gene expression can be completed, and the correlation between core gene expression and survival time of gastric cancer patients can be explored^26^. The ’ limma ’ R software package, ’ survival ’ R software package and ’ survminer ’ R software package were used to calculate the correlation between core gene expression and survival time and survival status of patients, and to explore the correlation between core gene expression and overall survival (OS), progression-free survival (FP) and post-progression survival (PPS) of patients with gastric adenocarcinoma.

### 2.11 Tumor microenvironment analysis

The " Estimate " R software package was used to complete the scoring of each gastric cancer sample in the TCGA database. The gastric cancer samples were divided into core gene high expression group and core gene low expression group according to the expression of core genes. StromalScore, ImmuneScore and ESTIMATEScore determined the effect of core gene expression on tumor microenvironment.

### 2.12 Tumor immune cell infiltration analysis

The ’ CIBERSORT ’ R package was used to calculate the degree of immune cell infiltration in each sample and to analyze the correlation between the expression of core genes and the degree of immune cell infiltration.

### 2.13 Immune checkpoint inhibitor expression analysis

The expression information of CD40, NRP1, TNFSF4, TNFRSF8, CD86, TNFRSF25, TNFRSF4, VTCN1, CD28, TNFSF18, TNFSF15, CD200, PDCD1LG2, LGALS9, TNFSF9, LAIR1, TNFSF14, ADORA2A and CD276 in gastric cancer was obtained, and the correlation between the expression of these immune checkpoint inhibitor genes and the core expression was analyzed.

### 2.14 Correlation analysis of tumor mutation burden

Tumor mutation burden (TMB) refers to the total number of somatic gene coding errors, base substitutions, gene insertions or deletion errors (mut / Mb) detected per million bases. High TMB tumor cells help anti-tumor T cell proliferation and anti-tumor response. It is currently an FDA-certified immunotherapy biomarker ^27^. The TMB data of gastric cancer samples in TCGA database were obtained, and the correlation between core gene expression and TMB was calculated.

### 2.15 Correlation between core gene expression and IPS

Immunophenoscore (IPS), as an indicator of tumor immunogenicity, can predict the effect of immunotherapy in cancer patients^28^. Immunotherapy data were obtained from The Cancer Immunome Atlas (https://tcia.at/home) to analyze the correlation between the expression of core genes and IPS treated with CTLA4 (-) PD1 (-), CTLA4 (-) PD1 (+), CTLA4 (+) PD1 (-) and CTLA4 (+) PD1 (+) in January 2023.

### 2.16 Small interfering RNA silencing core gene expression

The gastric cancer cells AGS and MKN45 were cultured in a 37 °C constant temperature cell incubator. In strict accordance with the instructions, 300,000 cells and six-well plates were inoculated respectively and cultured for 24 hours to make them fully adherent. When the cell concentration reached 80 %, it was taken out from the cell incubator. The siRNA was diluted according to the ratio of 50 ul Opti-MEM medium, 250 pmol siRNA and 12.5 ul TransIntro @ EL transfection reagent, and the siRNA-TransIntro @ EL complex was prepared by gently blowing and mixing. After 20 minutes at room temperature, the siRNA-TransIntro @ EL complex was added to a six-well plate with a cell density of 80 %. The cells were cultured in a 37 °C / 5 % constant temperature cell incubator. After 4 hours, the medium was replaced to improve the transfection efficiency. The transfection efficiency was observed by fluorescence microscopy at 24 hours and 48 hours. The cell protein was extracted, and the expression of the target protein was detected to further confirm the transfection efficiency, to select the siRNA with the highest transfection efficiency.

### 2.17 Statistics

Student *t* test was performed on two sets of data with normal distribution and homogeneity of variance, *t* ’ test was performed on data with normal distribution and heterogeneity of variance, and Mann-Whitney test was used for data that did not conform to normal distribution. The limma algorithm was used to calculate the differential genes in the transcriptome. The correlation between the expression of the target gene and the tumor microenvironment score was analyzed by Wilcox test. The correlation between the target gene and immune cell infiltration, immune checkpoint inhibitor gene expression and tumor mutation load was analyzed by Spearman test.

## Results

### 1 Viability curve of gastric cancer cells

CCK8 analysis showed that the growth of AGS cells and MKN45 cells was significantly inhibited after treatment with 0μM, 5μM, 12.5μM, 25μM, 50μM and 100μM cisplatin for 12 hours (Fig.1.A-B) and 24 hours (Fig.1.C-D). The IC50 value of AGS cells treated with cisplatin for 12 hours was 8.147μM. The IC50 value of MKN45 cells was 8.660μM.

**Figure 1:**
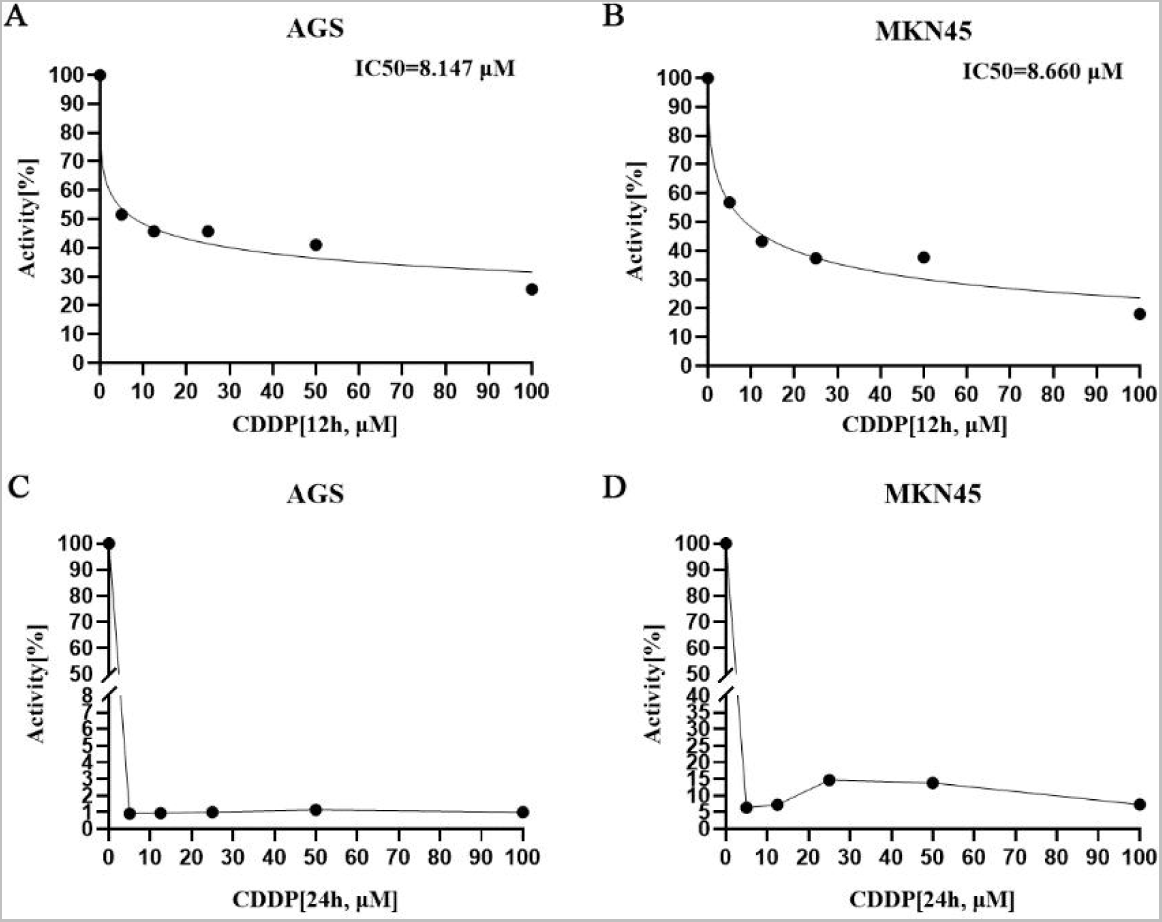
The cell viability curves of AGS cells (A) and MKN45 cells (B) co-cultured with IC50 concentration of cisplatin for 12 hours, and the cell viability curves of AGS cells (C) and MKN45 cells (D) cultured with IC50 concentration of cisplatin for 24 hours.

### 2 Identification of differential genes

The second-generation sequencing results showed that 9978 genes were significantly differentially expressed after MKN45 cells were treated with cisplatin 8.660 μM (IC50). Among them, 10 pyroptosis core genes NLRP6 (logFC = 3.270586096, p = 0.000708546), IL6 (logFC = 1.646344676, p = 0.000325408), GSDME (logFC = 0.666460514, p = 1.47581E-06), NLRP1 (logFC = 2.19681335, p = 9.2746E-15), CHMP6 (logFC = 1.27873683, p = 1.55749E-07), GSDMD (logFC = 0.742823512, p = 0.000133279), NLRC5 (logFC = 1.246191075, p = 1.20937E-19), TP53 (logFC = 1.860FC503494, p = 2.3254E-0.74823). Two genes CASP3 (logFC = − 2.226428178, p = 1.27397E-75) and TNFRSF10B (logFC = − 0.602568568, p = 1.25251E-07) were significantly decreased after cisplatin stimulation (Fig.2.A).

**Figure 2:**
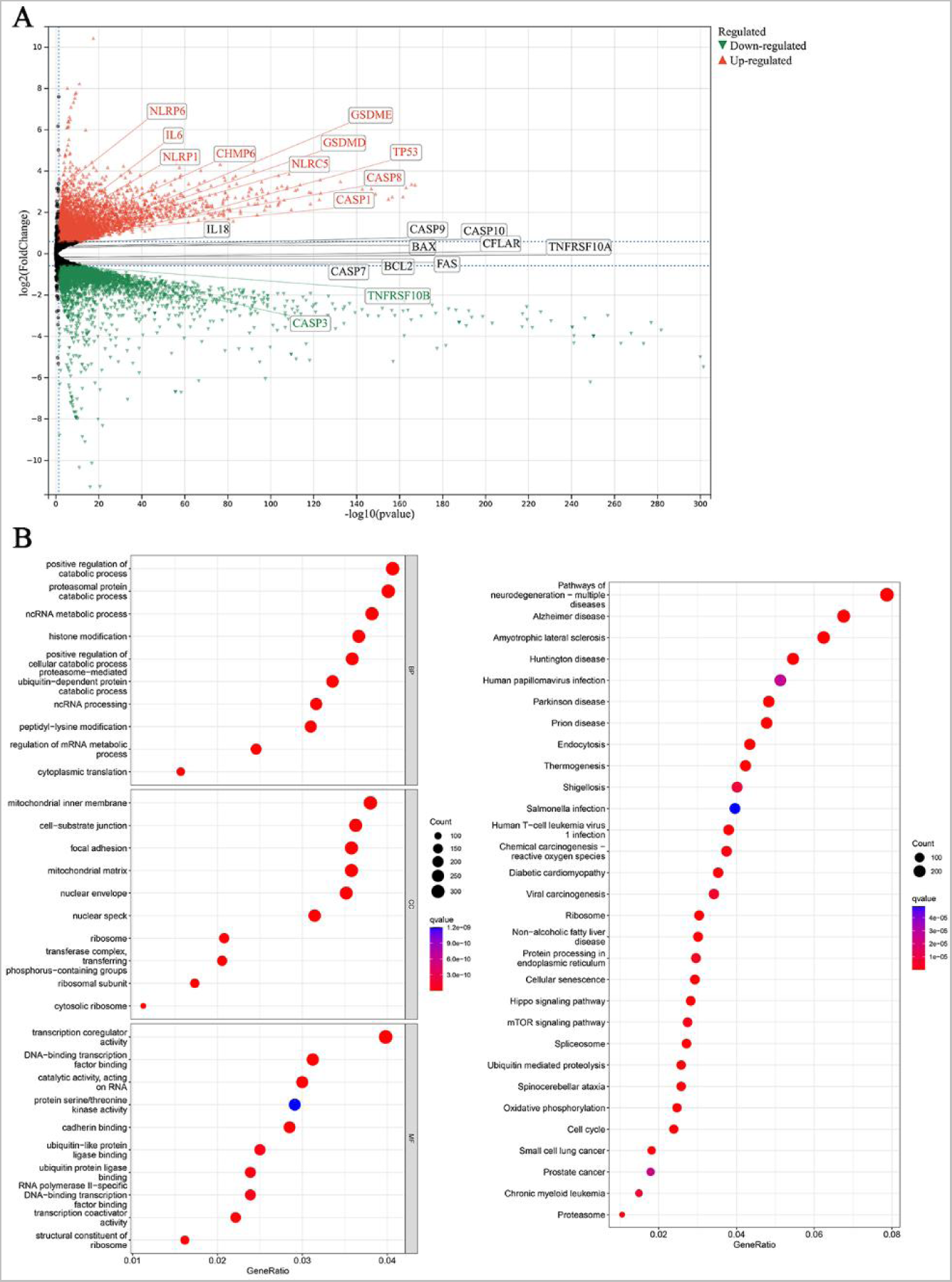
Volcano map based on differential genes after cisplatin treatment of MKN45 cells for 12 hours (A); after 12 hours of cisplatin treatment of MKN45 cells, GO enrichment analysis and KEGG enrichment analysis based on differential genes (B).

### 3 Gene enrichment analysis

GO enrichment analysis of differential genes before and after cisplatin treatment of MKN45 showed that differential genes were mainly concentrated in biological processes such as positive regulation of catabolic process, proteasome protein catabolic process and ncRNA metabolic process. Differential genes were mainly concentrated in cell component such as mitochondrial inner membrane, cell-substrate junction and focal adhesion. Differential genes were mainly concentrated in molecular function such as transcription coregulatory activity, DNA-binding transcription factor binding and catalytic activity, acting on RNA. Differential genes were mainly concentrated in pathways such as pathway of neurodegeneration-multiple diseases, Alzheimer disease, and Amyotrophic lateral sclerosis (Fig.2.B).

### 4 Cox regression analysis

The above ten pyroptosis genes significantly increased in MKN45 cells after 12 h of IC50 cisplatin stimulation were used to explore the effect of these ten genes on the overall survival of gastric cancer patients using Cox regression model. The results showed that GSDME was an independent prognostic factor for gastric cancer patients (Fig.3.A). CASP1, CASP8, CHMP6, GSDMD, IL6, NLRC5, NLRP1, NLRP6 and TP53 are not independent prognostic factors for gastric cancer patients (Fig.3.B-J).

**Figure 3:**
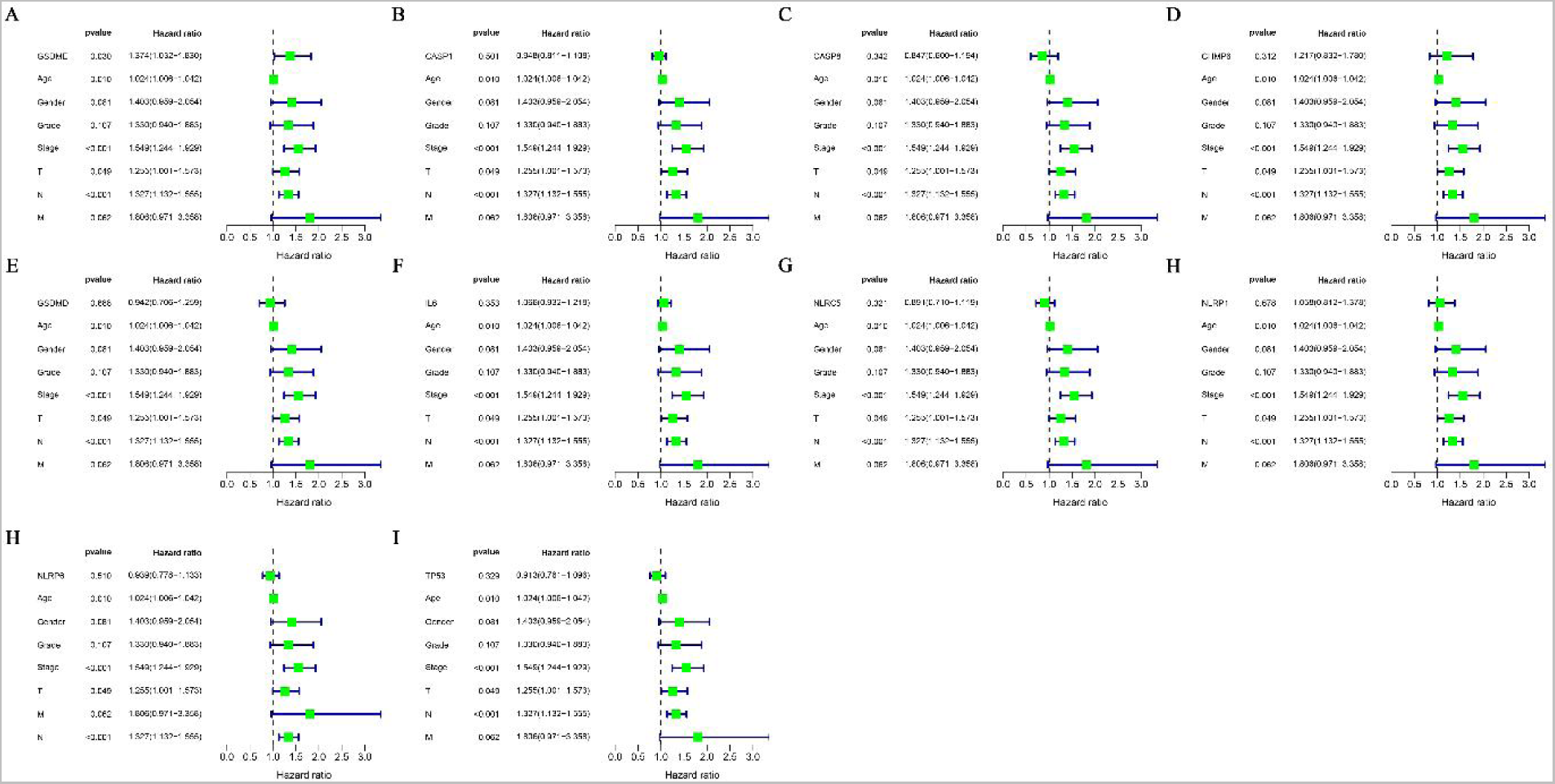
Single factor Cox regression analysis of pyroptosis core genes GSDME (A), CASP1 (B), CASP8 (C), CHMP6 (D), GSDMD (E), IL6 (F), NLRC5 (G), NLRP1 (H), NLRP6 (I) and TP53 (J) highly expressed in gastric cancer cells after cisplatin treatment.

**Figure 4:**
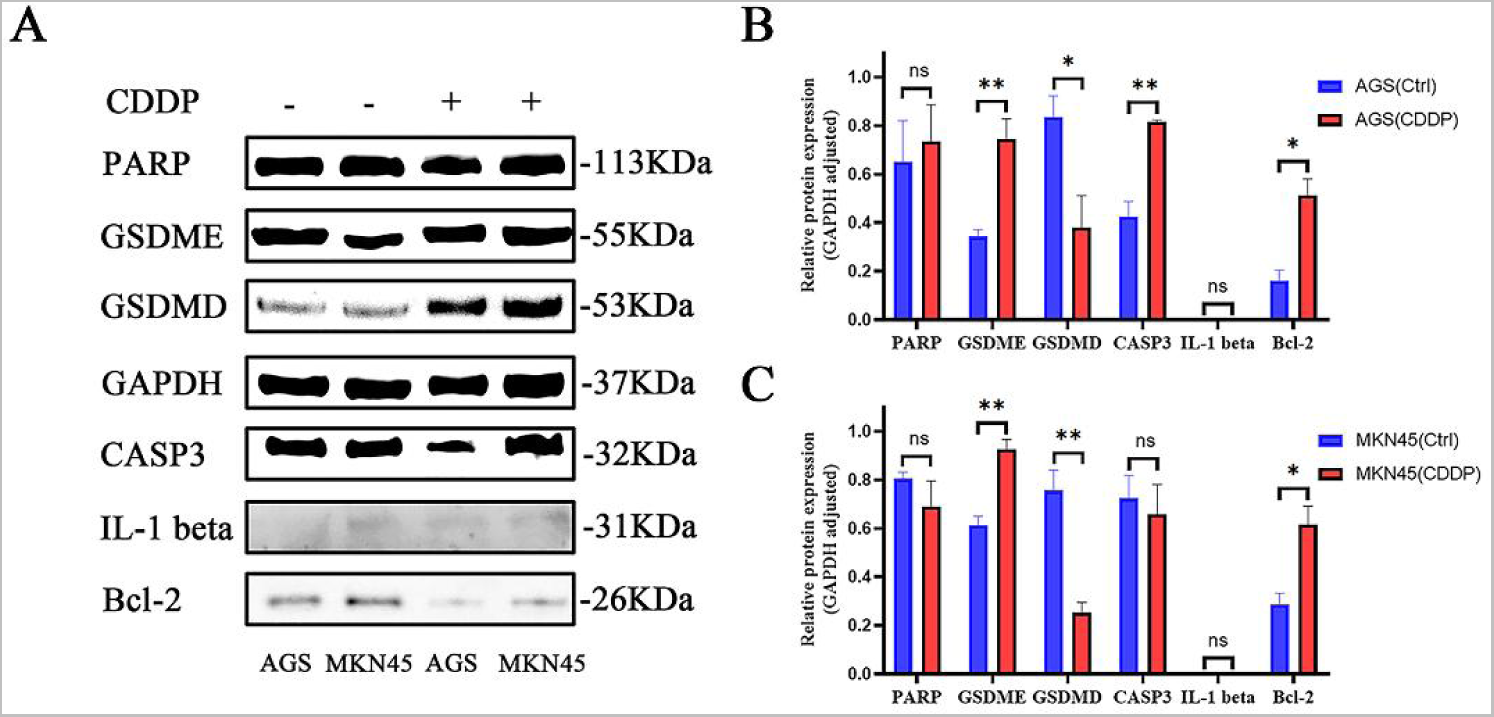
The changes of PARP, GSDME, GSDMD, GAPDH, CASP3, IL-1 beta, and Bcl-2 protein in gastric cancer cells after cisplatin treatment (A), and the statistical calculation results of expression in AGS cells (B) and MKN45 (C) cells, respectively.

### 5 Related changes after cisplatin treatment of gastric cancer cells

Western blot was used to detect the expression of GSDME protein (p = 0.0096), B-Cell CLL / Lymphoma 2 (Bcl-2) protein (p = 0.0124) and Caspase-3 (CASP3) protein (p = 0.0032) in human gastric adenocarcinoma AGS cells after cisplatin stimulation. The expression of GSDMD protein (p = 0.0455) was significantly decreased. There was no significant difference in the expression of Poly (ADP-Ribose) Polymerase (PARP) protein and Interleukin-1beta (IL-1 beta) protein. The expression of GSDME protein (p = 0.0047) and Bcl-2 protein (p = 0.0217) in human gastric cancer cell MKN45 showed a significant high expression trend, and the expression of GSDMD protein (p = 0.0061) also showed a significant decrease, while the expression of CASP3, PARP protein and IL-1 beta protein was not significantly different (Figure 5.A-C).

**Figure 5:**
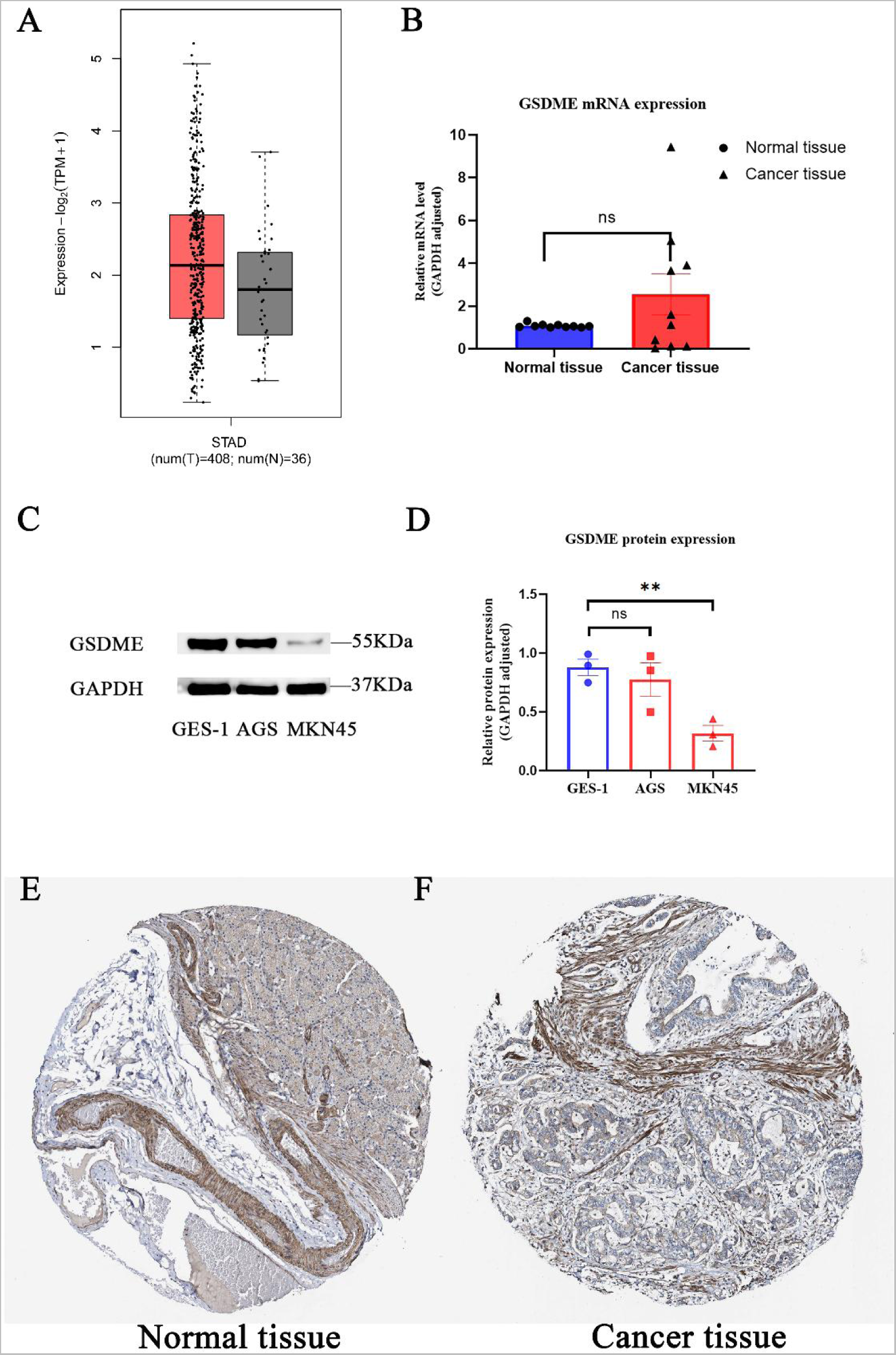
Differential expression of GSDME mRNA in gastric cancer tissues and adjacent tissues from TCGA database (A) and gastric cancer tissues (B); differential expression of GSDME protein between gastric cancer cells and normal gastric epithelial cells (C) and statistical calculation results (D); the expression of GSDME protein in normal gastric tissue (E) and gastric cancer tissue (F) was from HPA database.

### 6 Expression of GSDME in gastric cancer

The results showed that the expression of GSDME in gastric cancer tissues and adjacent tissues was statistically different (p = 0.0239), but the expression difference logFC was 0.3 (Fig.5.A). According to the results of qRT-PCR analysis, there was no significant difference in the expression of GSDME in 10 pairs of gastric cancer tissues and adjacent tissues (Fig.5.B). Western Blot analysis showed that compared with the expression of GSDME protein in gastric epithelial cells GES-1, there was no difference in the expression of human gastric adenocarcinoma cells AGS, but lower expression in MKN45 cells (Figure.5.C-D). Immunohistochemical images of GSDME in gastric cancer patients were collected from the human protein atlas database. The results showed that the expression of GSDME protein in gastric cancer tissues and adjacent tissues was mainly ’medium’ intensity (Fig.5.E-F).

### 7 Prognostic analysis

KM prognostic analysis showed that high expression of GSDME (DFNA5) was significantly associated with shorter OS in gastric cancer patients (p = 1.6e − 06, HR : 1.53 (1.28 − 1.82), Fig.6.A), and high expression of GSDME was significantly associated with shorter FP in gastric cancer patients (p = 1.7e − 05, HR : 1.55 (1.27 − 1.89), Fig.6.B). High GSDME expression was significantly associated with shorter PPS in gastric cancer patients (p = 0.00016, HR : 1.52 (1.22 − 1.9), Fig.6.C).

**Figure 6:**
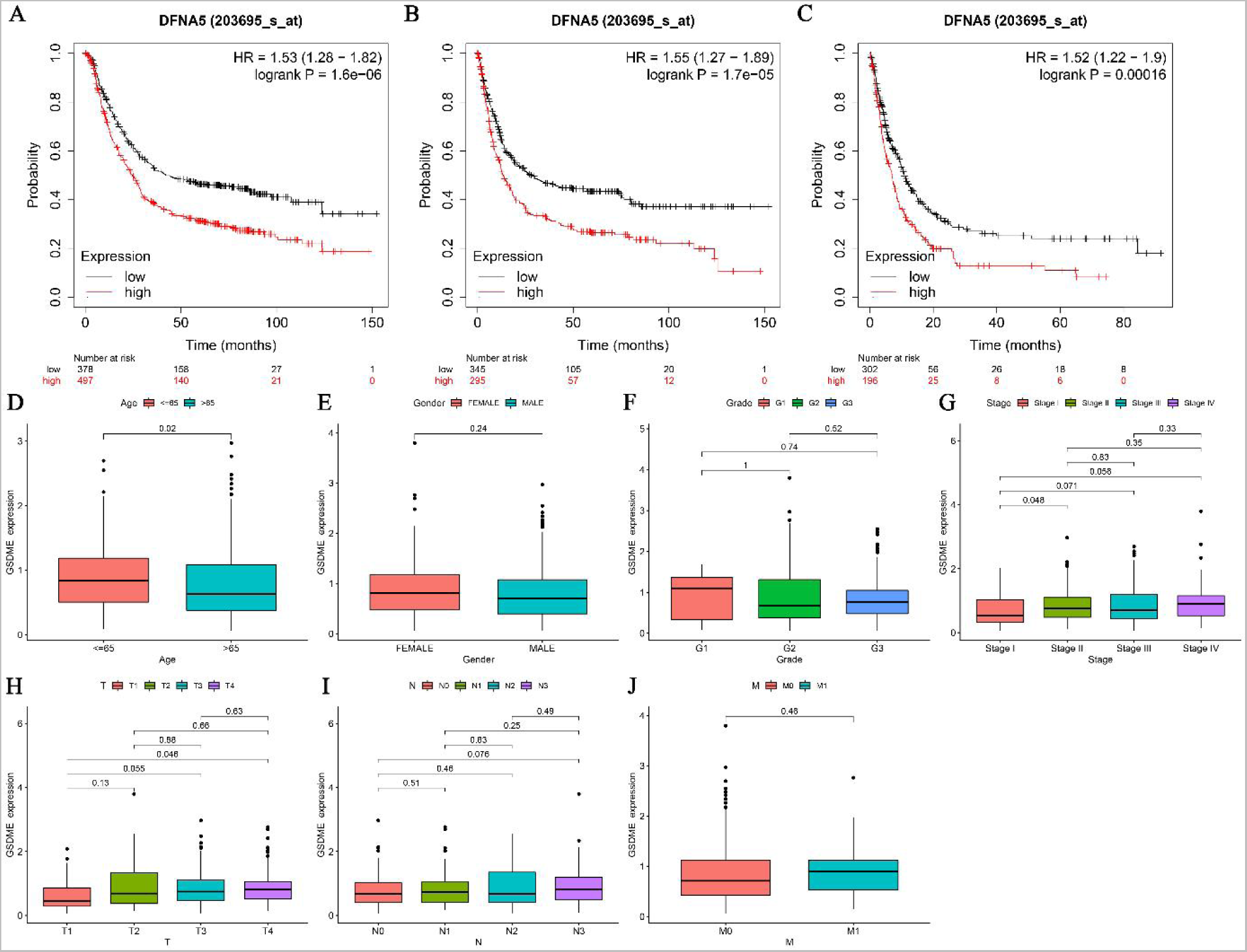
Correlation between GSDME expression and overall survival (A), progression-free survival (B) and post-progression survival (C) in patients with gastric cancer; the expression of GSDME in different age (D), gender (E), grade (F), stage (G), T stage (H), N stage (I) and M stage (J) of gastric cancer patients.

### 8 Correlation between GSDME and clinical characteristics of gastric cancer patients

The clinical analysis results showed that GSDME was highly expressed in gastric cancer patients less than or equal to 65 years old compared with patients older than 65 years old (Fig.6.D). Compared with Stage I stage, GSDME was higher expressed in Stage II patients (Fig.6.G) ; compared with T1 stage, GSDME had higher expression in T4 patients (Fig.7.H). There was no significant difference in the expression of GSDME in gastric cancer patients with different genders (Fig.6.E), different Grades (Fig.6.F), different N stages (Fig.6.I) and different M stages (Fig.6.J).

**Figure 7:**
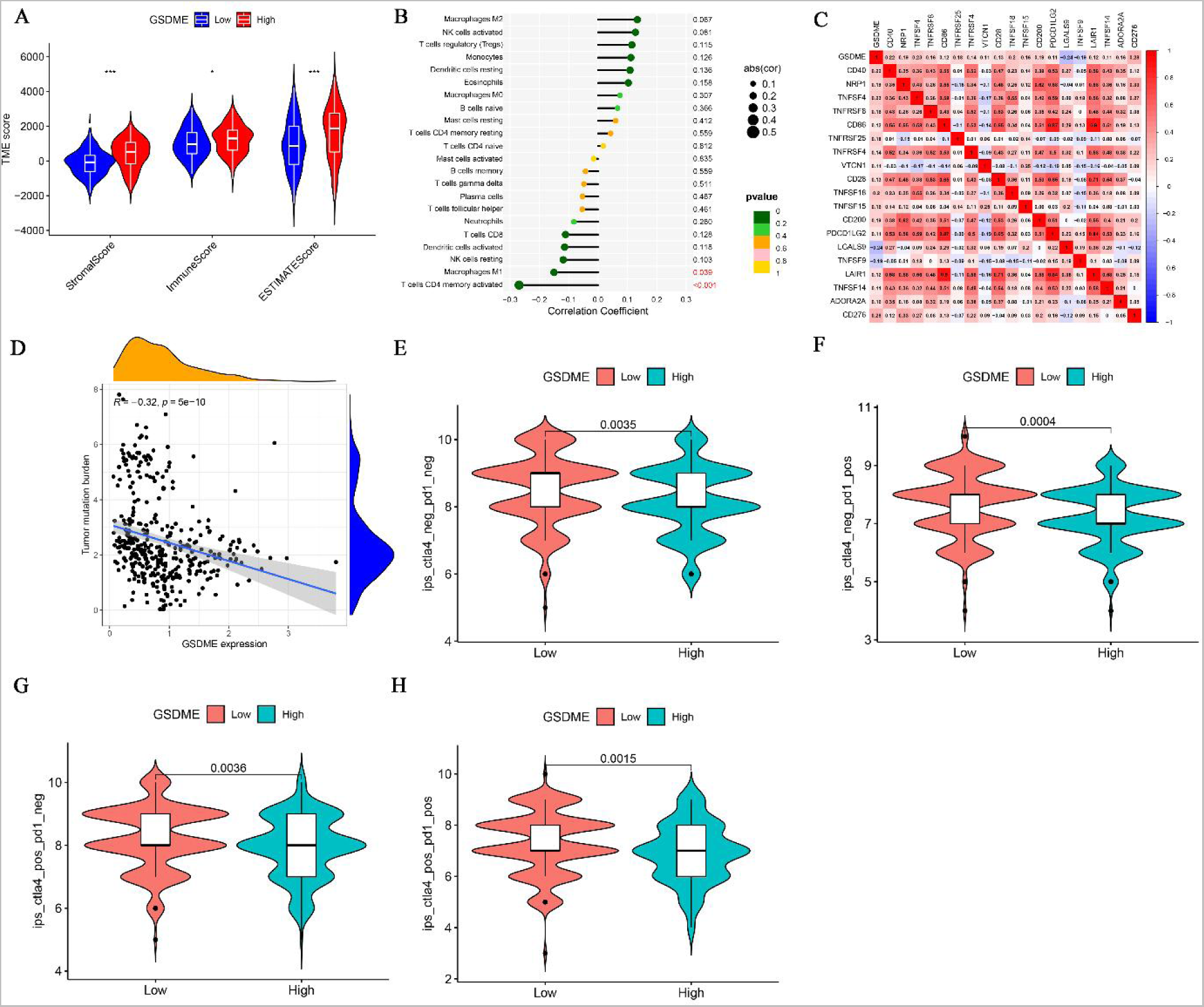
Correlation between GSDME expression and StromalScore, ImmuneScore, and ESTIMATEScore in tumor microenvironment (A), and correlation between GSDME and immune cell infiltration (B); the correlation between GSDME expression and immune checkpoint inhibitor gene expression in gastric cancer (C); the expression of GSDME in gastric cancer was correlated with tumor mutation burden (D); the correlation between GSDME expression in gastric cancer and IPS(F-H) treated with CTLA4 (-) PD1 (-), CTLA4 (-) PD1 (+), CTLA4 (+) PD1 (-) and CTLA4 (+) PD1 (+).

### 10 Correlation between GSDME expression and tumor microenvironment score

The results of immune microenvironment analysis showed that the expression of GSDME was significantly positively correlated with StromalScore, ImmuneScore and ESTIMATEScore (Fig.7.A).

### 11 Correlation between GSDME expression and immune cell infiltration

The immune cell infiltration analysis results showed that the expression of GSDME was significantly positively correlated with Macrophage M1 and T cells CD4 memory activated (Fig.7.B).

### 12 Correlation between GSDME expression and immune checkpoint inhibitor expression

In gastric cancer, the expression of GSDME was positively correlated with the expression of CD40, NRP1, TNFSF4, TNFRSF8, CD86, TNFRSF25, TNFRSF4, VTCN1, CD28, TNFSF18, TNFSF15, CD200, PDCD1LG2, LAIR1, TNFSF14, ADORA2A and CD276, but negatively correlated with the expression of LGALS9 and TNFSF9 (Fig.7.C).

### 13 Correlation between GSDME expression and TMB

TMB analysis showed that GSDME expression was significantly negatively correlated with tumor mutation burden in gastric cancer (Fig.7.D).

### 14 Correlation between GSDME expression and IPS

Immunotherapy analysis showed that the expression of GSDME was significantly negatively correlated with IPS in patients (Fig.7.E-H) with gastric cancer treated with CTLA4 (+) PD 1 (+), CTLA4 (+) PD 1 (-) and CTLA4 (-) PD 1 (+).

### 15 Changes after transfection of siRNA GSDME

Compared with NC siRNA treatment, AGS cell treated with GSDME small interfering RNA had significantly better proliferation ability at 12 hours (p = 0.0031) and 24 hours (p = 0.0104) after gastric cancer cells were treated with GSDME small interfering RNA (Fig.8.A) ; MKN45 cell had significantly worse proliferation ability at 24 hours (p < 0.0001) and 48 hours (p = 0.0339) (Fig.8.B). The results of Western Blot experiments and analysis showed that after GSDME expression was silenced, the expression of PARP protein (p = 0.0259) and Bcl-2 protein (p = 0.0022) in human gastric adenocarcinoma AGS cell decreased significantly, while the expression of CASP3 protein (p = 0.0239) increased significantly, and the expression of GSDMD protein (p = 0.7022) and IL-1 beta protein (p = 0.6126) did not change significantly. The expression of Bcl-2 protein (p = 0.0353) was significantly decreased in human gastric cancer cell line MKN45, while the expression of CASP3 protein (p = 0.0136) was significantly increased. The expression of PARP protein (p = 0.1922), GSDMD protein (p = 0.3427) and IL-1 beta protein (p = 0.7945) did not change significantly (Fig.8.C-E).

**Figure 8:**
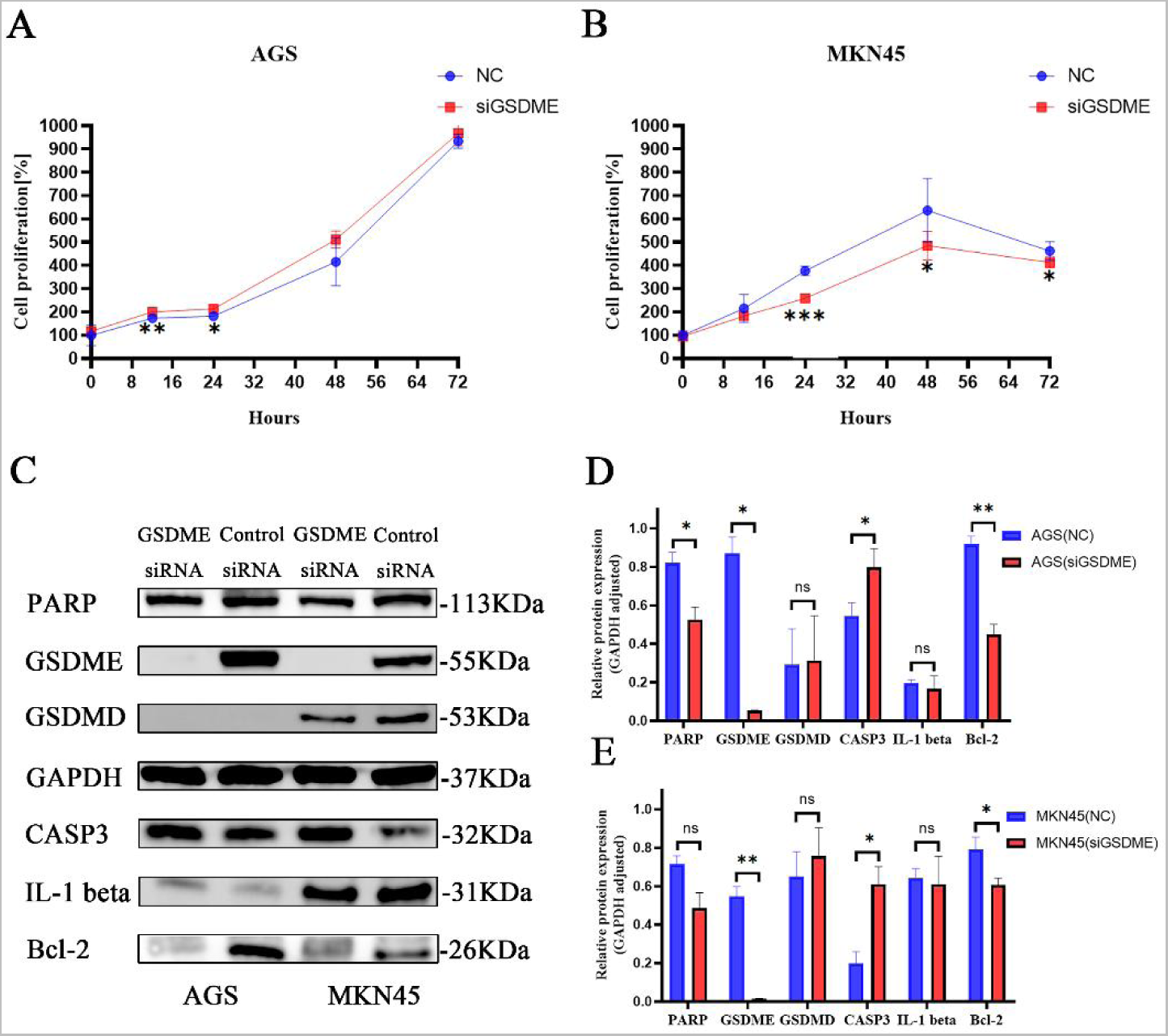
Changes in the proliferation of AGS cells (A) and MKN45 (B) cells after silencing the expression of GSDME by siRNA; the changes of PARP, GSDMD, GAPDH, CASP3, IL-1 beta, and Bcl-2 proteins after silencing the expression of GSDME by siRNA (C), and the difference analysis results in AGS cells (D) and MKN45 cells (E).

### 16 The effect of GSDME expression on cisplatin cytotoxicity

CCK8 results analysis showed that when the expression of GSDME in AGS cells and MKN45 cells was silenced by siRNA, the cytotoxicity of cisplatin at 12 h was significantly reduced, and the survival rate of AGS cells (p < 0.001) and MKN45 (p < 0.001) cells treated with cisplatin at 12 h IC50 was significantly increased (Fig.9.A). Western Blot results showed that after cisplatin treatment of gastric cancer cells with IC50 concentration, compared with NC siRNA gastric cancer cells, in addition to the low expression of GSDME protein in AGS cells of siGSDME, GSDMD protein (p = 0.0034), IL-1 beta protein (p = 0.0024) and Bcl-2 protein (p = 0.0468) showed significantly low expression, CASP3 protein (p = 0.0020) showed significantly high expression; in addition to the low expression of GSDME protein in siGSDME MKN45 cells, GSDMD protein (p = 0.0099), IL-1 beta protein (p = 0.0002) and Bcl-2 protein (p = 0.0102) showed significantly low expression, while CASP3 protein (p = 0.0256) showed significantly high expression (Fig.9.B-D).

**Figure 9:**
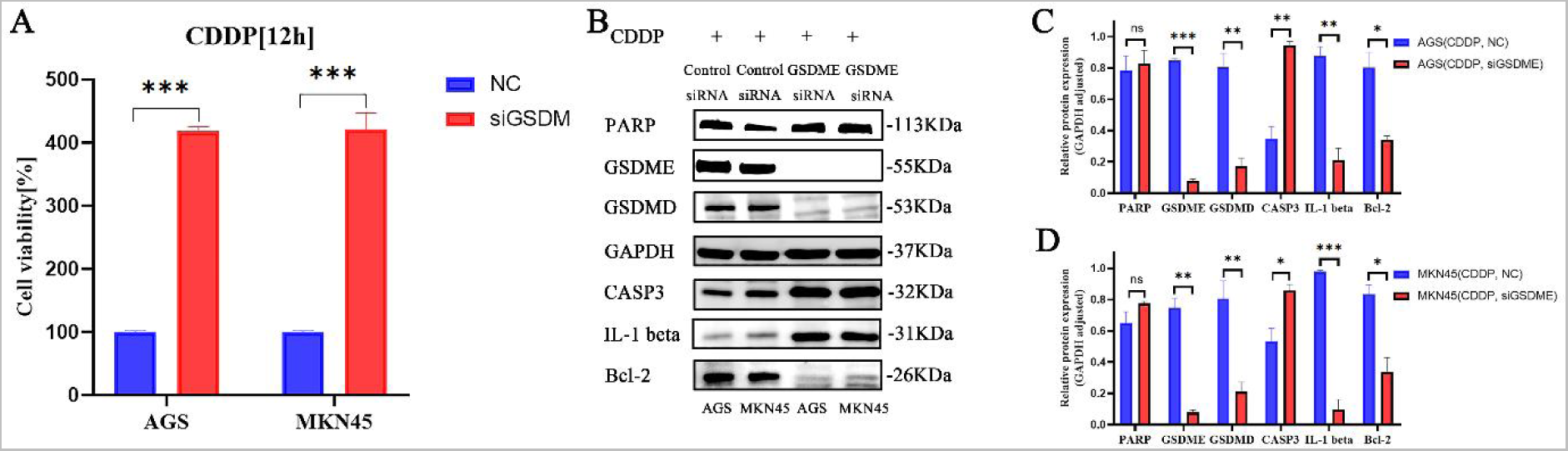
Changes in cell viability (A), PARP, GSDMD, GAPDH, CASP3, IL-1 beta, and Bcl-2 proteins in gastric cancer cells treated with cisplatin after silencing GSDME by siRNA (B) and the calculation results in AGS cells (C) and MKN45 (D) cells.

## Discussion

Cisplatin is widely used in chemotherapy for cancer, germ cell tumors, lymphoma and sarcoma. The traditional view is that cisplatin interacts with purine bases and interferes with intracellular DNA repair, resulting in irreparable DNA damage, which mediates apoptosis. In this study, the second-generation sequencing results showed that most apoptotic core genes did not show significant high expression after cisplatin treatment of human gastric cancer cell MKN45. Pyroptosis-related genes (NLRP6, IL6, GSDME, NLRP1, GSDMD, NLRC5, CASP8 and CASP1) showed a significant high expression trend after cisplatin treatment, indicating that cisplatin may promote the death of gastric cancer cells by activating pyroptosis. In 2001, Souza et al.first proposed the term ’ pyroptosis ’ to distinguish between apoptosis and pyroptosis, which belong to programmed cell death^29^. The study of shao et al.showed that GSDME was specifically cleaved by CASP3 in the adaptor under the action of chemotherapeutic drugs, resulting in GSDME-N fragment, which eventually led to pyroptosis of cancer cells expressing GSDME^15^. Cisplatin can activate pyroptosis through the MEG3 / NLRP3 / caspase-1 / GSDMD pathway to exert anti-tumor effects in triple-negative breast cancer^21^. Similarly, cisplatin can also mediate pyroptosis in esophageal cancer cells and lung cancer cells^23, 30^. In this study, the enrichment analysis of differential genes before and after cisplatin treatment showed that the differential genes were mainly concentrated in neurodegenerative diseases, amyotrophic lateral sclerosis, Alzheimer ’s disease, Parkinson ’s disease, human papillomavirus infection and shigellosis. Current research has found that abnormal activation of programmed cell death, including pyroptosis, leads to unnecessary loss of neuronal cells and functions, which is one of the main characteristics of neurodegenerative diseases such as amyotrophic lateral sclerosis, Alzheimer ’s disease, Parkinson ’s disease and Huntington ’s disease^31, 32^. Pyroptosis is closely related to Alzheimer ’s disease. The activation of inflammasome can promote the accumulation of amyloid protein and the deterioration of neuronal function in APP / PS1 mice, leading to the onset of Alzheimer ’s disease^33^. Using drugs to target the occurrence of pyroptosis mediated by NLRP3 inflammasome can improve the condition of Alzheimer ’s disease and Parkinson ’s disease^34, 35^. Shigella is the pathogen of bacillary dysentery. There is a clear relationship between Shigella and the occurrence of pyroptosis. It can inhibit the activation of inflammasomes in epithelial cells, optimize the interaction with the host and establish a successful infection by regulating the ’open’ and ’close’ of inflammasomes^36^. The differential genes after cisplatin treatment of gastric cancer cells are concentrated in the signaling pathways of pyroptosis-related diseases, which also shows that there may be a strong correlation between cisplatin and pyroptosis.

The second-generation sequencing results showed that GSDME was significantly highly expressed after cisplatin treatment of cancer cells, and Western Blot also showed that GSDME protein was significantly highly expressed after cisplatin treatment of gastric cancer cells. GSDME protein is an independent prognostic factor in patients with gastric cancer, and previous studies have also shown that GSDME protein is an executive protein of chemotherapeutic drug-mediated pyroptosis^37^. Therefore, GSDME was selected as the core gene of cisplatin acting on gastric cancer cells for further analysis. The results of prognostic analysis showed that the high expression of GSDME was a risk factor for gastric cancer patients, which was significantly correlated with shorter OS, FP and PPS in gastric cancer patients. GSDME has the potential to be a prognostic marker for gastric cancer. Studies have shown that prediction models constructed using pyroptosis-related genes have good predictive performance in the diagnosis or prognosis of gastric cancer or immunotherapy^38–41^. Through KEGG enrichment analysis based on GESA, it was found that GSDME was negatively correlated with signal pathways such as aminoacyl-tRNA biosynthesis, DNA replication and nucleotide excision repair in gastric cancer. GSDME may inhibit DNA repair behavior in gastric cancer, which is also consistent with the related cognition of pyroptosis-mediated cancer cell death. Pyroptosis is a double-edged sword for tumor. The relationship between pyroptosis and tumor environment exists in various ways: it can kill cancer cells and promote inflammation to affect the formation and development of tumor^42, 43^. After silencing GSDME with siRNA, different cells showed different growth and proliferation. GSDME-silenced AGS cells showed stronger proliferation ability after 12 hours and 24 hours of culture, while GSDME-silenced MKN45 cells showed worse proliferation ability after 24 hours and 48 hours of culture. However, after 72 hours GSDME-silenced AGS cells and MKN45 cells showed the same proliferation ability as NC group AGS cells and MKN45 cells. Cells with different proliferation ability previously showed the same proliferation ability after 72 hours, which may be due to the timeliness of siRNA. However, other studies have also shown that GSDME may not be directly involved in the development of tumors, and the expression of GSDME protein is not always directly related to the volume and weight of tumors, but may play a key role in tumor immunity and chemotherapy-mediated cell death^44, 45^.

GSDME was significantly positively correlated with StromalScore, ImmuneScore and ESTIMATEScore in tumor microenvironment, and also significantly positively correlated with Macrophage M1 and T cells CD4 memory activated. GSDME was positively correlated with TMB in patients with gastric cancer, and negatively correlated with IPS in patients receiving CTLA4 (+) PD 1 (+), CTLA4 (+) PD 1 (-), CTLA4 (-) PD 1 (+) and CTLA4 (-) PD (-) immunotherapy. These results indicate that GSDME affects the tumor immunity of gastric cancer, and the high expression of GSDME may indicate that it is difficult for gastric cancer patients to achieve ideal results in immunotherapy. Although GSDME is difficult to be used as a diagnostic marker for gastric cancer, it can potentially be used as a marker for gastric cancer immunotherapy. Recent studies have shown that although GSDME ablation cannot protect cells from final cell death, its protein expression may still play a role in tumor immunogenicity, which can induce (secondary) necrotic cell death to enhance immunogenicity^46^. This provides another evidence for GSDME as an immunotherapy marker. The increase in immunogenicity after GSDME cleavage may also be why the expression of GSDME is significantly positively correlated with TMB and the expression of most immune checkpoint inhibitor genes. For cancer cells with normal expression of GSDME, they showed CASP3-dependent GSDME activation after chemotherapy treatment^15, 47–49^. The toxicity of cisplatin on gastric cancer cells decreased after GSDME silencing, which indicated that the sensitivity of chemotherapeutic drugs was directly related to the expression of GSDME, and the decrease of GSDME expression affected the efficiency of chemotherapy. Previous studies have also shown that after GSDME is silenced, A-549 cells induce less cell death due to cisplatin^48^. The decrease of cisplatin sensitivity in clinical practice may be affected by the expression of core genes such as GSDME. Some studies have found that knockout of GSDME transforms platinum-induced cell death from pyroptosis to apoptosis^45^. The results of Western Blot in this study showed that the silencing of GSDME in gastric cancer cells significantly increased the expression of CASP3 regardless of the effect of cisplatin. This indicates that CASP3 may mediate the inhibition of GSDME expression. This also indicates that there is also a CASP3-GSDME mutual regulation / feedback mechanism in gastric cancer cells^50^. The results showed that the expression of GSDME protein was synergistic with the expression of Bcl-2 protein in gastric cancer cells. After cisplatin treatment, GSDME protein and Bcl-2 protein were highly expressed in gastric cancer cells. A decrease in Bcl-2 expression also accompanied the silencing of GSDME. Bcl-2 protein can inhibit the apoptosis induced by radiotherapy, chemotherapy and other tumor treatments, and the up-regulation of Bcl-2 is related to the resistance of various cancer cell lines to cisplatin^51, 52^. However, the expression of Bcl-2 is a good prognostic marker for gastric cancer patients in Asian countries^53^. The expression of GSDME, an executive protein of pyroptosis, is an unfavorable factor for the prognosis of gastric cancer patients. Further study on the regulatory system of GSDME and Bcl-2 is helpful to reveal the mystery of pyroptosis as a ’double-edged sword’ for the occurrence and development of cancer. Chemotherapy drugs including cisplatin have side effects such as nausea and vomiting, acute kidney injury, neurotoxicity and ototoxicity during use^54^. Studies have shown that acute kidney injury is significantly related to pyroptosis, and baicalin can alleviate acute kidney injury by regulating the ROS / NLRP3 / CASP1 / GSDMD signaling pathway^55^. Currently, most of the studies on ear protection are only achieved by limiting cisplatin-induced apoptosis. Studies have shown that mutations in GSDME, TJP2 and MSRB3 can lead to single-gene hearing impairment^56^. Cisplatin may mediate the occurrence of pyroptosis by activating GSDME, resulting in ototoxicity. The in-depth study of pyroptosis in ototoxicity and the executive protein GSDME may be conducive to the discovery of more effective methods for ear protection. The results showed that after cisplatin treatment of gastric cancer cells, GSDMD and GSDME were significantly up-regulated at the transcriptome and protein levels. The upregulation of GSDMD protein was significantly associated with ocular hypertension / glaucoma, lung injury in mice, mastitis in mice, cardiac hypertrophy, chicken cardiotoxicity and neurotoxicity^57–62^. GSDME, an executive protein mediated by chemotherapy drugs, is indispensable in killing cancer cells. Without affecting the expression of GSDME, inhibiting the expression of GSDMD protein seems helpful to solve the side effects caused by chemotherapy drugs, but further research is needed to clarify its feasibility. This study is mainly based on cell experiments and has not completed in vivo experimental verification; no clinical trials have been carried out to further prove the clinical value of GSDME. However, in general, based on various research methods, we have a new understanding of the mechanism of cisplatin on gastric cancer cells, and also found that GSDME, a core factor of pyroptosis, has the potential to be a prognostic marker and immunotherapy marker for gastric cancer.

Pyroptosis as an innate immune mechanism mediates the death of tumor cells. On the other hand, as a way of pro-inflammatory cell death, it may also provide a suitable environment for tumor growth. In-depth study of the mechanism of pyroptosis and the precise regulation of pyroptosis are helpful to improve the killing efficiency of cancer cells and reduce the side effects of drugs.

## Conclusion

In this study, we identified that the pyroptosis-related genes were highly expressed after cisplatin treatment of gastric cancer cells. The pyroptosis core gene GSDME was an independent prognostic factor for gastric cancer patients. There was no significant difference in the expression of GSDME between gastric cancer tissues and adjacent tissues. The silencing of GSDME significantly reduced the sensitivity of AGS cell and MKN45 cell to cisplatin. The expression of GSDME was significantly negatively correlated with TMB and IPS after immunotherapy. GSDME can potentially be a biomarker for predicting the sensitivity of chemotherapy, immunotherapy and prognosis in patients with gastric cancer.

## Declarations

### Ethics approval and consent to participate

This study was approved by the Medical Ethics Committee of Gansu Provincial People ’s Hospital (ID: 2022-316), Lanzhou, China. All patients signed the informed consent and approved the study.

### Consent for publication

Not applicable

### Data Availability Statement

The datasets analyzed during the current study are available in TCGA (https://portal.gdc.cancer.gov/), Gene Expression Profiling Interactive Analysis 2 (http://gepia2.cancer-pku.cn/), Kaplan Meier plotter portal (https://kmplot.com/analysis/), GEO database (https://www.ncbi.nlm.nih.gov/), The Molecular Signatures Database (https://www.gsea-msigdb.org/gsea/msigdb), and The Human Protein Atlas (https://www.proteinatlas.org/).

### Author Contributions

Xianglai Jiang and Yongfeng Wang conceived the study, Chenyu Wang completed the data collation and processing, Miao Yu provided the support of methodology and Haizhong Ma and Hui Cai completed the work of Funding acquisition. Xianglai Jiang completed the draft, Hui Cai reviewed the paper.

## Abbreviation

GSDME: Gasdermin E
GSDMD: Gasdermin D
IL-6: Interleukin-6
CASP1: caspase-1
IC50: Half inhibitory concentration
TCGA: The Cancer Genome Atlas Program
STAD: Stomach adenocarcinoma
GO: Gene Ontology
KEGG: Kyoto Encyclopedia of Genes and Genomes
OS: overall survival
FP: free progression
PPS: post progression survival
IPS: Immunophenoscore
Bcl-2: B-Cell CLL/Lymphoma 2
CASP3: caspase-3
PARP: Poly(ADP-Ribose) Polymerase
IL-1 beta: Interleukin-1beta

## Acknowledgments

This work has benefited from aforementioned databases. The authors thank the Key Laboratory of Molecular Diagnostics and Precision Medicine for Surgical Oncology in Gansu Province for their help and support in the methodology.

